# Application of ARIMA and Holt-Winters forecasting model to predict the spreading of COVID-19 for India and its states

**DOI:** 10.1101/2020.07.14.20153908

**Authors:** Mrutyunjaya Panda

## Abstract

The novel Corona-virus (COVID-2019) epidemic has posed a global threat to human life and society. The whole world is working relentlessly to find some solutions to fight against this deadly virus to reduce the number of deaths. Strategic planning with predictive modelling and short term forecasting for analyzing the situations based on the worldwide available data allow us to realize the future exponential behaviour of the COVID-19 disease. Time series forecasting plays a vital role in developing an efficient forecasting model for a future prediction about the spread of this contagious disease. In this paper, the ARIMA (Auto regressive integrated moving average) and Holt-Winters time series exponential smoothing are used to develop an efficient 20-days ahead short-term forecast model to predict the effect of COVID-19 epidemic. The modelling and forecasting are done with the publicly available dataset from Kaggle as a perspective to India and its five states such as Odisha, Delhi, Maharashtra, Andhra Pradesh and West Bengal. The model is assessed with correlogram, ADF test, AIC and RMSE to understand the accuracy of the proposed forecasting model.

## 1. Introduction

Ever since the discovery of COVID-19 epidemic that originated in Wuhan, China in late 2019, lots of clinical investigations are going on by World Health Organization (WHO) and other medical practitioners/companies across the globe to develop vaccines to countermeasures this disease. This has tremendously affected human life with illness, economic slowdown and death. Looking into the impact of this deadly disease over more than 210 countries, recently WHO has declared COVID-19 pandemic [1] as a global threat to public health and social life and advised some preventive strategies to be followed strictly including: wearing of masks, maintaining social distancing of 3 feet, washing of hands by sanitizer repeatedly, avoid moving in large public gatherings etc., till some well-tested drugs are being discovered to counter this virus. COVID-19, being SARS Cov-2 causes severe respiratory problems where the patient needs emergency care with ICU facility and in many cases with high mortality rate. The present clinical experience after using infected patient’s swab test and abnormal chest CT imaging suggests Lungs CT imaging may be the better diagnostic method in place of nucleic acid testing for early detection of such a contagious disease [2]. It is observed that SARS Cov-2 COVID-19 spread more rapidly among human through personal contact in comparison to other related respiratory diseases. More importantly, around 20% of the COVID-19 infected humans do not have any symptoms [3]. It is also observed that Cov-2 is more dangerous to elderly people aged above 60 years and to those having of past disease history with weak immunity, cardiac, cancer, HIV/AIDS and diabetic etc. [4].

Looking into all these, there is an emergent need to minimize the negative impact of COVID-19, raid processing, trend analysis and future forecasting of relevant data in terms of death and recovered cases. As of today, most of the countries perform manual testing using testing kits and expensive blood test, which is a time-consuming process demand for improvement in the medical testing process. To improve this manual testing process, machine learning may be seen as an alternative solution efficient and effective predictive treatment of the disease [5-7].

### Motivation and Objectives

Being the second-most populous country in the world, India has fought well to contain the COVID-19 spread by having a mere one in million infected cases as per the recent status available, motivated us to carry out this research to develop some forecast models for future prediction.

ARIMA and Holt-Winters methods mark their suitability in forecasting the COVID-10 disease spread with the number of daily as well as cumulative confirm and death cases, for better decision making to tackle the situation beforehand.

### The objectives of the paper are

1. To find the effect of COVID-19 epidemic as a perspective of India and its five states in terms of confirmed and death cases.
2. To develop predictive models using ARIMA and Holt-Winters additive time series with a 95% confidence interval (CI)
3. To perform 20-days ahead forecasting of COVID-19 pandemic using Holt-Winters and ARIMA prediction models using AIC and Root mean square error (RMSE)
4. To compare with official value with our predicted value for their effectiveness in the decision-making process

After presenting an introduction in Section 1, Section 2 presents some of the COVID-19 related work available in the literature to date. Section 3 presents the materials and methods used in this research. Experimental results, performance evaluation and discussion are provided in Section 4, followed by conclusion in section 5.

## 2. Related Work

Machine learning has found its enormous application across various public health including disease prediction and relevant valid drug development [3]. Rough set theory considered to be an effective method to deal with health care data having inconsistent and imprecise information [8]. Machine learning and deep learning is applied successfully in medical imaging applications, cancer tumour classification and tuberculosis (TB) disease prediction and analysis [9-12].

In a recent review, Gamboa [13] discusses the usefulness of various stochastic models such as AR, ARIMA and GARCH etc. and then mentioned the scarce applications of deep learning in time series forecasting, even though the use of neural networks for financial prediction is not new [14]. Deep learning is applied in computerized tomography scan and radiogram images of the COVID-19 patients to detect the presence of viral infections [15] with good performance accuracy.

Ribeiro et al. [16] performed extensive applications of short-term forecasting techniques in Ten Brazilian States using ARIMA, random forest, Support vector regression (SVR), RIDGE and an ensemble approach to perform one to six days ahead forecasting on cumulative confirmed COVID-19 patients and ranked the model performance based on having low mean absolute error.

Tuli et al. [17] use a robust iterative weighting model to statistically predict the severity of COVID-19 spread efficiently and compared with the baseline Gaussian model and advocates that a poorly fitting model could lead to worsening the public health situation.

Ghosal et al. [18] uses autoregression, multiple regression and linear regression techniques for trend analysis of COVID-19 death patients at the 5^th^ and 6^th^ week in India and opines autoregression for better prediction performance.

Kumar, Gupta and Srivastava [19] presents a progressive search considering the latest modern approaches available that might be suitable to fight with COVID-19 pandemic and opines for the further research as technology contribution to reduce the impact of this outburst.

Deb and Majumdar [20] explored the usefulness of time series forecasting for trend analysis at an early stage of the COVID-19 epidemic for different countries for developing countermeasure policies in dealing with the epidemic.

Kucharski et al. [21] discuss the early spread pattern of COVID-19 in every nook and cranny of China with different available datasets using scientific methods and explore its possible spread in other parts of China.

Dey et al. [22] use a visual method to understand the COVID-19 disease patterns everywhere possible in the world, for better containment planning.

## 3. Materials and Methods

### COVID-19 Dataset

The publicly available time-series data is obtained from Kaggle [23] that collects the data as and when released in the official website of the Ministry of Health and Family Welfare (MoHFH) Government of India. The MoFHW updates the national level as well as state-level confirmed, death and recovered data. This dataset consists of three parts as (i) complete list of all states arranged in day-wise details, (ii) national daily and cumulative details and (iii) state-wise daily details of confirmed, death and recovered cases. In all the three parts, data are collected from 30^th^ January to 29^th^ June 2020. The complete list contains a total of 3511 instances, the national part containing daily and cumulative cases with 153 instances and state-level data with 109 instances. The dataset has 10 attributes with time-stamp, state, latitude, longitude, confirmed cases, recovered cases. death cases and their new reported cases for each one, on a day to day basis. In the case of India, the first reported case was on 30^th^ Jan 2020 and the first reported death case was on 11^th^ March 2020.

### Holt-Winters Additive time series forecasting

The Holt-Winters forecasting algorithm developed by Charles Holt and Peter Winters is useful for time series forecasting where users smooth the time series data and then use it for the forecast as per its interest. Exponential smoothing is a method to smooth a time series where it allocates exponentially decreasing weights and values in opposition to historical data to lessen the value of the weights for the bygone data. Exponential smoothing can be classified into three types. While the simple or single exponential smoothing time series forecasting for uni-variate data does not have any systematic structure with no trend and seasonality. In this case, an only single parameter *α* is used as a smoothing factor that lies between 0 and 1. A smaller *α* value designate slow learning, takes more past observations for forecasting and a larger value designate faster learning takes most recent observations for making a forecast. Next type is double exponential smoothing where apart from *α*, another smoothing parameter *β* is used for change in trend. There are two types of the trend such as additive trend which gives linear trend analysis and the other is multiplicative trend gives exponential trend analysis. It is observed that during multi-step long-range forecasting, the trend may become unfeasible. Dampening may be practical hereby reducing the trend size for the future forecast with a straight line (no trend). Finally, triple exponential smoothing adds seasonality (*γ*) part from *α* and *β*. This is the most recent exponential smoothing method, named after its inventor Charles Holt and Peter Winters, which is useful to find the changing pattern of level, trend and seasonality over time by using either additive or multiplicative seasonality. In this paper, Holt-Winter additive exponential smoothing [24], is used for forecasting,

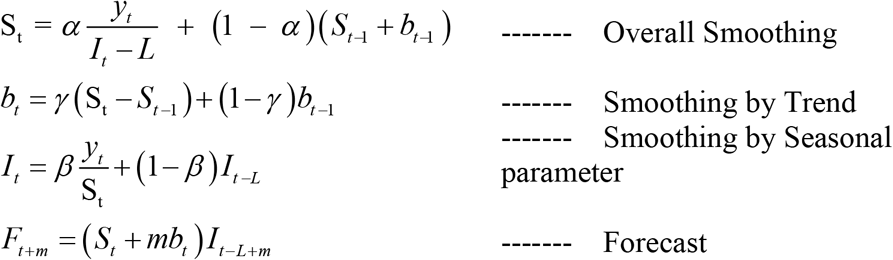

Where, α is the smoothing factor for the level; *β* is the smoothing factor for the trend, *γ* is the smoothing factor for the seasonality; y and S are actual and smoothed observation; b is the trend factor, I is the seasonal index, F is the forecast at *m* steps ahead, L is cycle length and t is a period. The value of*α, β* and *γ* are to be chosen carefully so that the error is minimized.

### ARIMA

The autoregressive integrated moving average (ARIMA) is a very popular and efficient time series forecasting model that can predict a value in time series by combining its past values and errors. The ARIMA was initially coined by Box and Jenkins and then named after their inventors.

The working principle of ARIMA lay down by three stages:

Step-1: Identification stage, where the input time series data are used for computing auto-correlation (ACF) and partial auto-correlation (PACF) from correlogram and auto-correlogram respectively, to check whether the data is stationary or non-stationary. Next, if the time series data is non-stationary, then it is to be converted to a stationary one using differencing. With differencing, one can model the change from one time period to the next rather than modelling the series itself. It is worth noting here that over differencing may increase standard deviation. Finally, the white noise residual test will enable us to perform hypothesis testing on the auto-correlation of the series for its statistical significance.

Step-2: Estimation stage with statistical analysis helps us to understand the adequacy of the model. This step presents the table of the goodness of fit statistic in terms of AIC (Alkaike Information Criterion) and BIC (Bayesian Information Criterion) for comparing the various models, with lowest is a better strategy in choosing the best model. Further, an outlier analysis is done to check whether there are any possible changes not yet incorporated in the estimated model. Sometimes, this seems a cumbersome task for a long series and hence, a post-hoc outlier analysis is advised in such a case.

Step-3: Forecasting stage with the help of estimation stage, could generate the best prediction output with future time series values along with upper and lower bound of these forecast with confidence interval using the best ARIMA model.

Generally, ARIMA can be modeled as ARIMA (p,d,q) with p is the order of AR (autoregressive) process, d is the differencing and q is the order of MA (moving average) part. If d=0, then ARIMA (p,d,q) reduces to an ARMA(p,q) model.

## 4. Experiments and Analysis

### 4.1. Modelling and evaluation

Predictive modelling with statistical and machine learning methods that takes the past data is used for making the better future prediction. To have a better prediction model, selection of most appropriate methodology is of paramount importance, failing which it may not only provide worst decision making but also a greater chance of affecting public health and social life. The forecasting model is a popular choice when it comes to deal with past numerical data and to predict the new value-based on past data. Short term forecasting model for time series data with short term predictions is considered in this paper using Holt-Winters additive and ARIMA architecture, to explore further insights to deal with COVID-19 epidemic in the perspective of India and five Indian states such as Odisha, Andhra Pradesh, Delhi, Maharashtra and West Bengal. The proposed methodology is shown in Figure 1.

**Figure 1:**
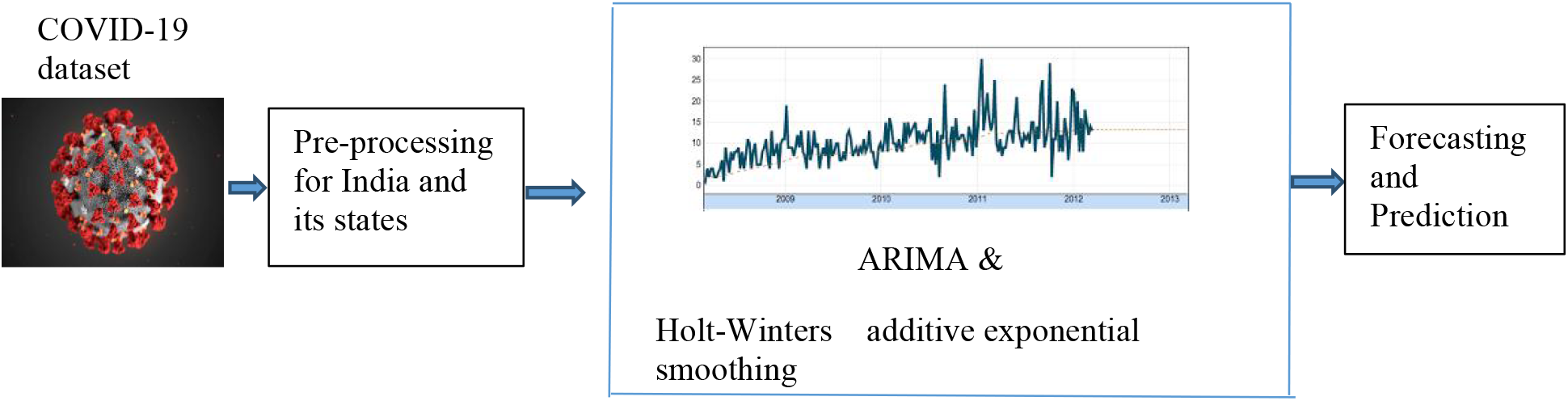
Proposed time series forecasting method

### 4.2. Parameter setting in the proposed forecasting methods

The open-source statistical software ‘R’ and various statistical and time series packages such as ‘Xrealstats and forecast sheet of Microsoft excel 2016’ are used for this research purpose.

### Experimental Results- India Forecasting

To make the COVID-19 time-series data to be stationary, ADF (Augmented Dickey-Fuller) Test is carried out with differencing d=0, 1 and 2 with the result shown in Table 1 and Table 2. It is seen that d=1 is a proper choice to make the series stationary at a 5% significance level test in comparison to d=0 with the low p-value.

**Table 1:** ADF test for confirming cases.

| *test* | *lag* | *stat* | *p-value* |
| --- | --- | --- | --- |
| bartlett | 1 | 0.035915 | 0.016678 |
| box-pierce | 1 | 4.527442 | 0.033355 |
| ljung-box | 1 | 4.531313 | 0.03328 |

**Table 2:** ADF test for Death cases.

| *test* | *lag* | *stat* | *p-value* |
| --- | --- | --- | --- |
| box-pierce | 1 | 5.904672 | 0.015101 |
| ljung-box | 1 | 5.90972 | 0.015058 |

Further, to estimate other two parameters of the model such as ACF and PACF; Correlogram and Partial Correlogram of the time series and first difference are used for lag 1 to lag 30, which is shown in Figure 2. The results ACF and PACF plots for confirming and death cases on time series COVID-19 data for India shows that there is a single spike in both cases at lag 1. All other coefficients for lag 2 to lag 30 are well within the significance limits for both ACF and PACF plots. Hence, the proposed ARIMA models with various alternatives are compared with their AIC values using Ljung-Box and box-pierce test. Finally, ARIMA (4,1,1) is selected for forecasting confirmed cases and ARIMA(3,1,1) is used for death prediction purpose, out of all the possible models with low AIC values and well-behaved residuals with zero mean and constant variance. The potential ARIMA models for final selection are shown in Table 3.

**Table 3:** ARIMA (p,q,d) modelling for confirmed and death cases with AIC values-India.

| case/model | 101 | 102 | 201 | 202 | 301 | 302 | 311 | 401 | 411 |
| --- | --- | --- | --- | --- | --- | --- | --- | --- | --- |
| confirmed | 67634 | - | 67237 | 69370 | 67030 | 68820 | - | 66844 | 65816 |
| death | 45040 | 47699 | 44702 | 46777 | 44507 | 46226 | 43463 | - | - |

**Figure 2:**
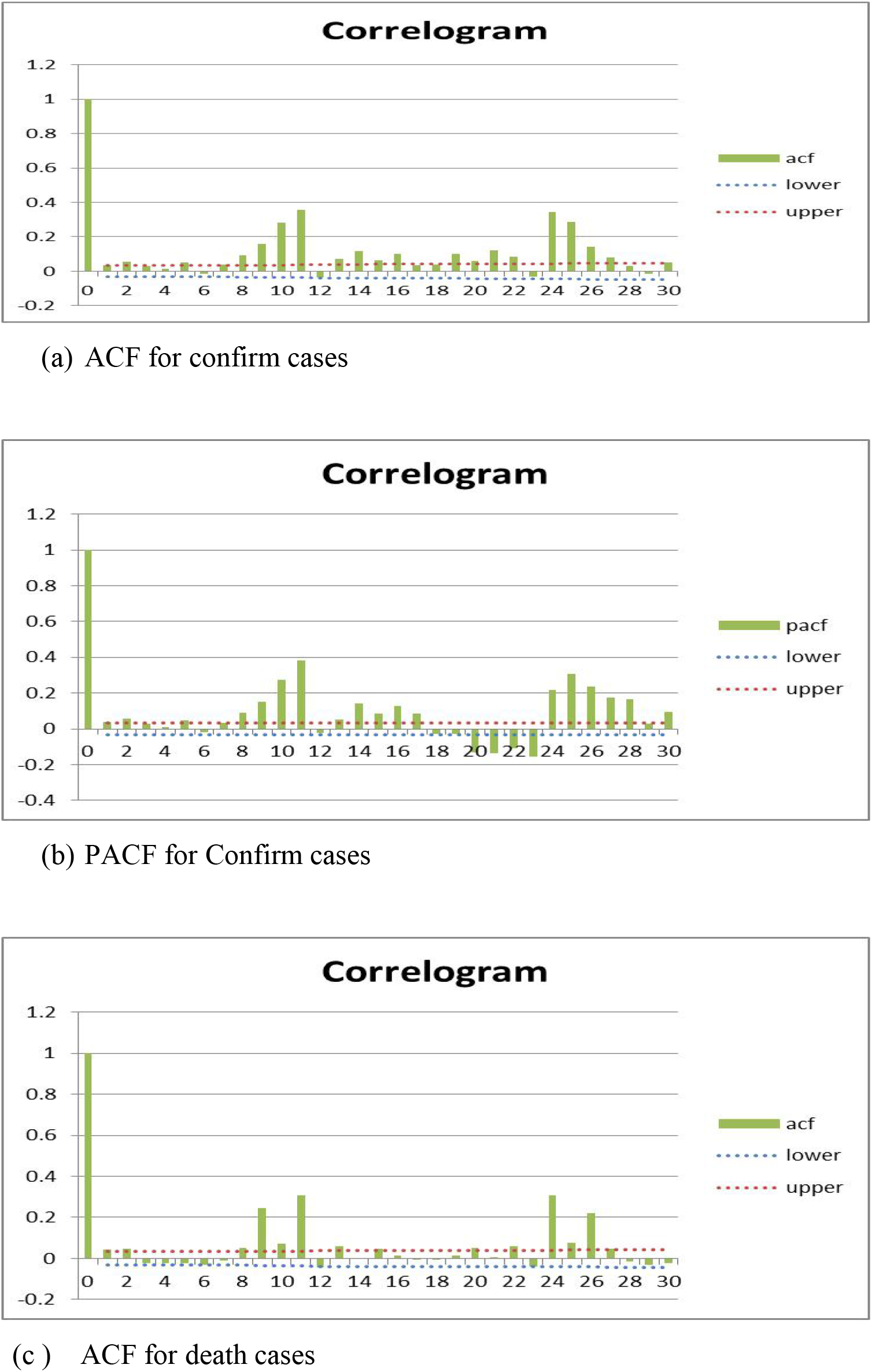
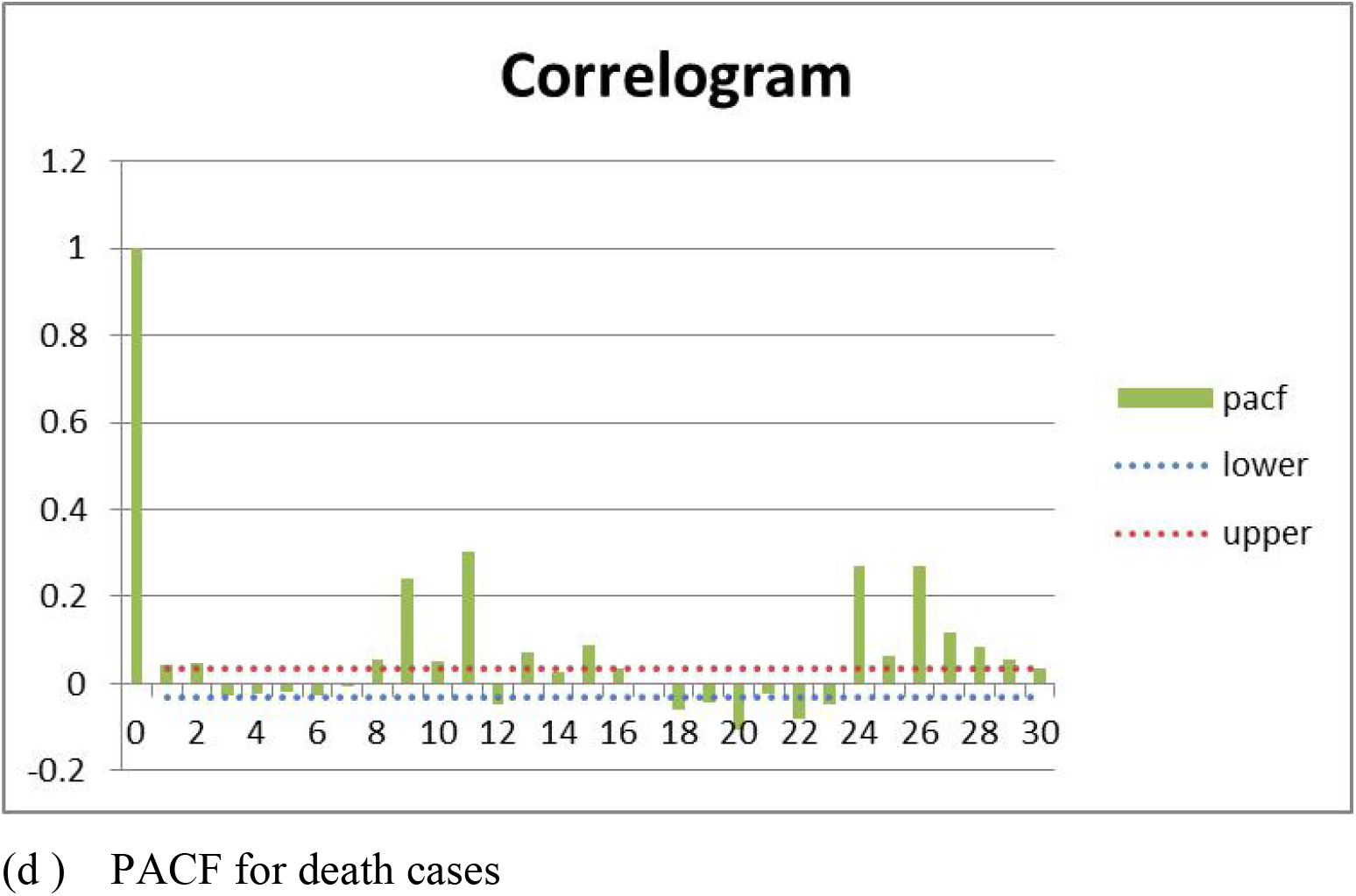
ACF and PACF for confirmed and death cases.

For national daily data, we get AIC with confirming cases in ARIMA-401 and 411 with 1862 and 1866 respectively. Similarly, the AIC value of 1501 and 1453 for ARIMA-301 and 311 for India daily death cases respectively. From these, it is evident that ARIMA-411 and ARIMA-311 is the most potent model with the lowest AIC value for predicting confirms and death cases for India as a whole.

Using ARIMA (4,1,1) prediction model, forecast result showing daily confirmed and ARIMA (3,1,1) showing death cases for India with 95% CI, up to August 31, 2020 are shown in Table 4. Further, to check the accuracy of the proposed forecasting model, a comparison has also been performed for the daily confirmed and death cases for India from 1^st^ July 2020 to 10^th^ July 2020, as shown in Table 5.

**Table 4:**
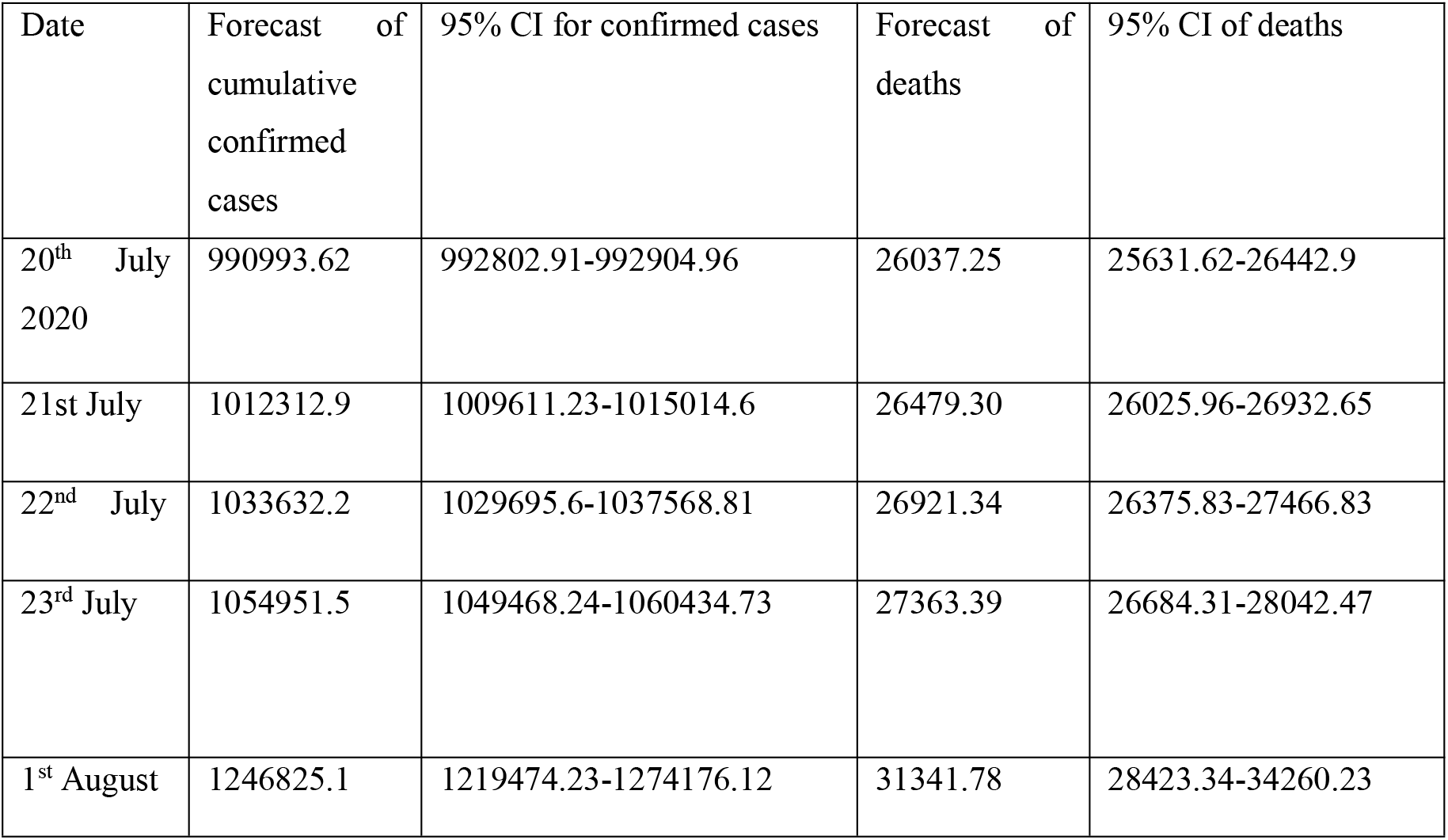
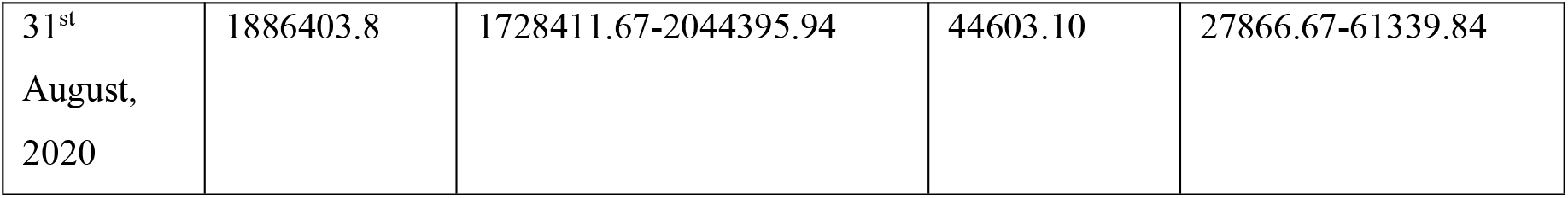
Results of forecasting for confirmed and death cases using the ARIMA model until 31^st^ August 2020-India.

| Date | Forecast of cumulative confirmed cases | 95% CI for confirmed cases | Forecast of deaths | 95% CI of deaths |
| --- | --- | --- | --- | --- |
| 20<sup>th</sup> July 2020 | 990993.62 | 992802.91-992904.96 | 26037.25 | 25631.62-26442.9 |
| 21<sup>st</sup> July | 1012312.9 | 1009611.23-1015014.6 | 26479.30 | 26025.96-26932.65 |
| 22<sup>nd</sup> July | 1033632.2 | 1029695.6-1037568.81 | 26921.34 | 26375.83-27466.83 |
| 23<sup>rd</sup> July | 1054951.5 | 1049468.24-1060434.73 | 27363.39 | 26684.31-28042.47 |
| 1<sup>st</sup> August | 1246825.1 | 1219474.23-1274176.12 | 31341.78 | 28423.34-34260.23 |
| 31<sup>st</sup><br>August,<br>2020 | 1886403.8 | 1728411.67-2044395.94 | 44603.10 | 27866.67-61339.84 |

**Table 5:**
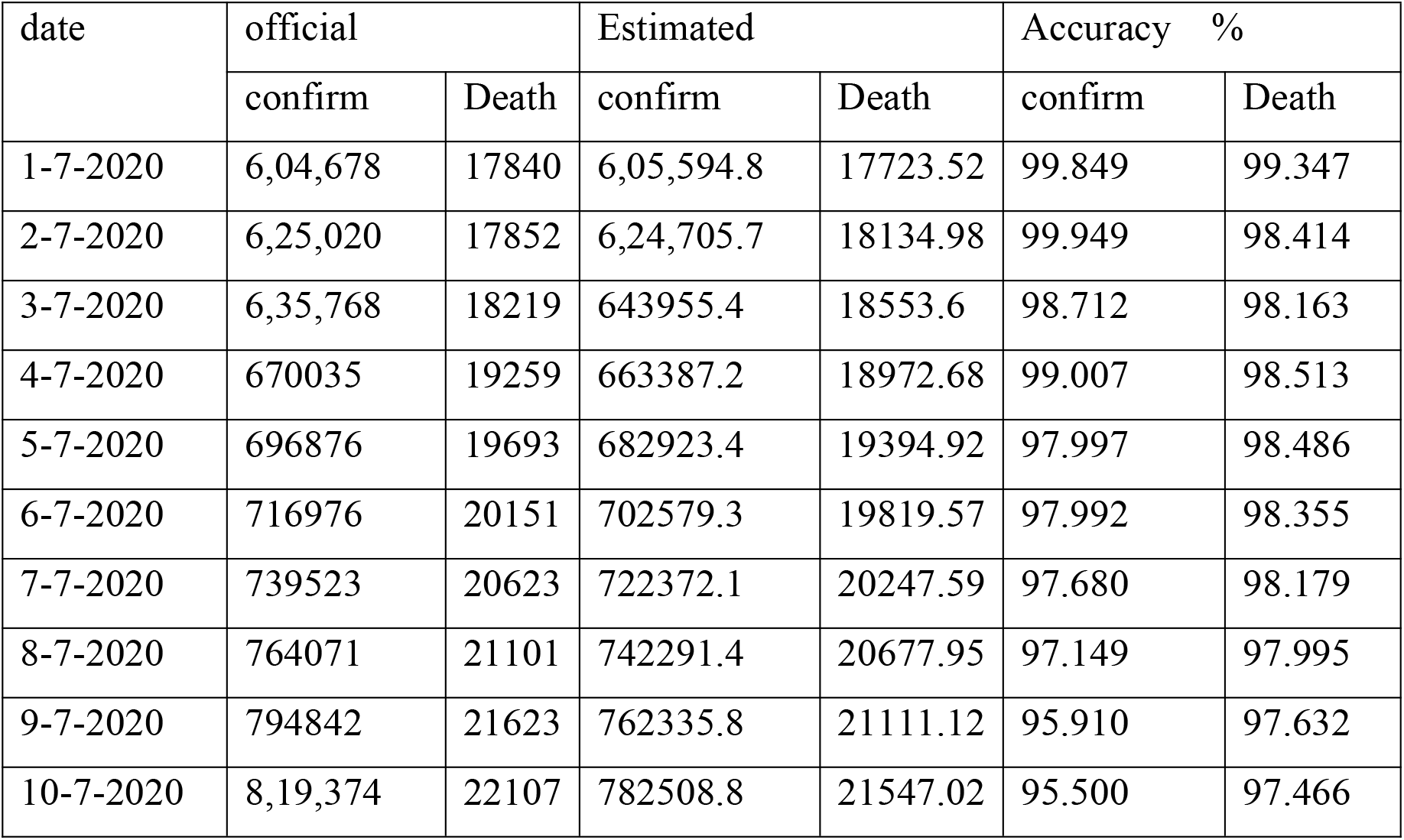
comparison of 10-days with official and estimated confirmed and death cases using ARIMA - India.

| date | official |  | Estimated |  | Accuracy % |  |
| --- | --- | --- | --- | --- | --- | --- |
|  | confirm | Death | confirm | Death | confirm | Death |
| 1-7-2020 | 6,04,678 | 17840 | 6,05,594.8 | 17723.52 | 99.849 | 99.347 |
| 2-7-2020 | 6,25,020 | 17852 | 6,24,705.7 | 18134.98 | 99.949 | 98.414 |
| 3-7-2020 | 6,35,768 | 18219 | 643955.4 | 18553.6 | 98.712 | 98.163 |
| 4-7-2020 | 670035 | 19259 | 663387.2 | 18972.68 | 99.007 | 98.513 |
| 5-7-2020 | 696876 | 19693 | 682923.4 | 19394.92 | 97.997 | 98.486 |
| 6-7-2020 | 716976 | 20151 | 702579.3 | 19819.57 | 97.992 | 98.355 |
| 7-7-2020 | 739523 | 20623 | 722372.1 | 20247.59 | 97.680 | 98.179 |
| 8-7-2020 | 764071 | 21101 | 742291.4 | 20677.95 | 97.149 | 97.995 |
| 9-7-2020 | 794842 | 21623 | 762335.8 | 21111.12 | 95.910 | 97.632 |
| 10-7-2020 | 8,19,374 | 22107 | 782508.8 | 21547.02 | 95.500 | 97.466 |

Looking into all the above estimations, short term forecasting is performed till August 31, 2020, for both confirmed and death cases, which are shown in Figure 3 and Figure 4.

**Figure 3:**
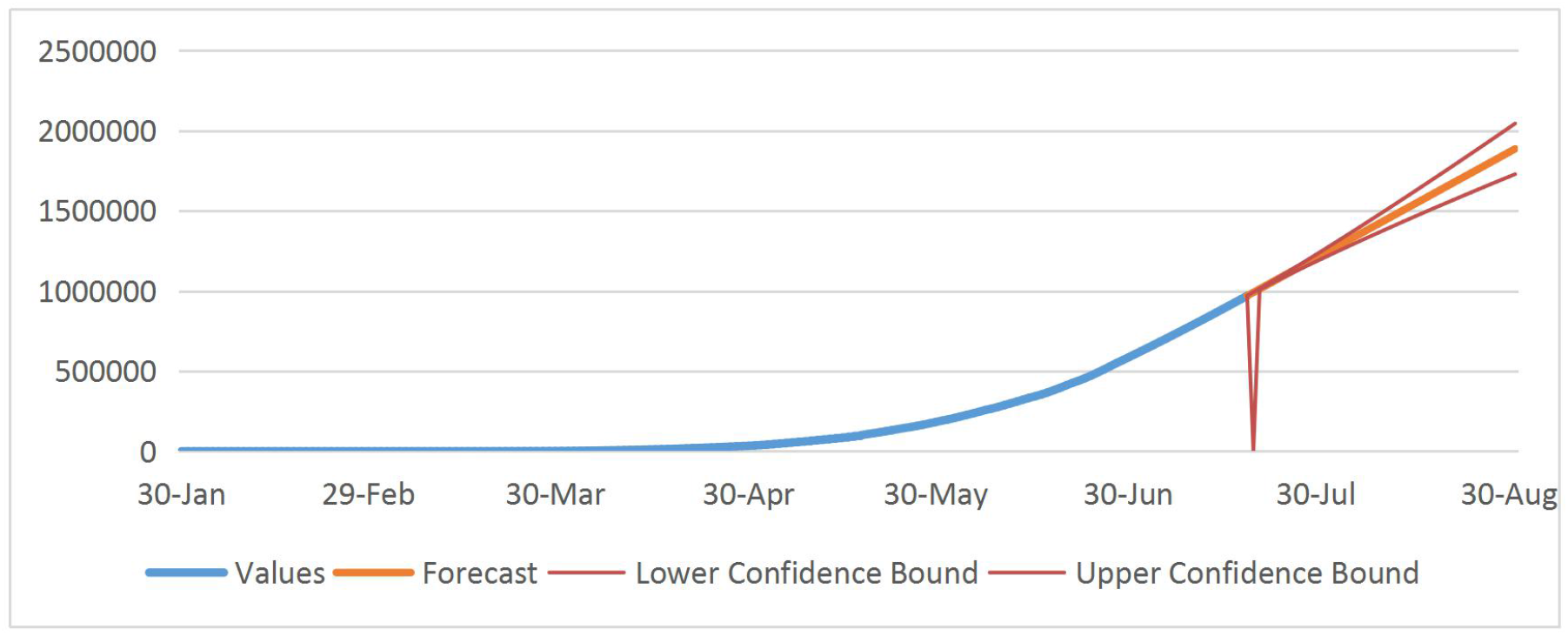
Forecasting for the confirmed cases (ARIMA-411): India

**Figure 4:**
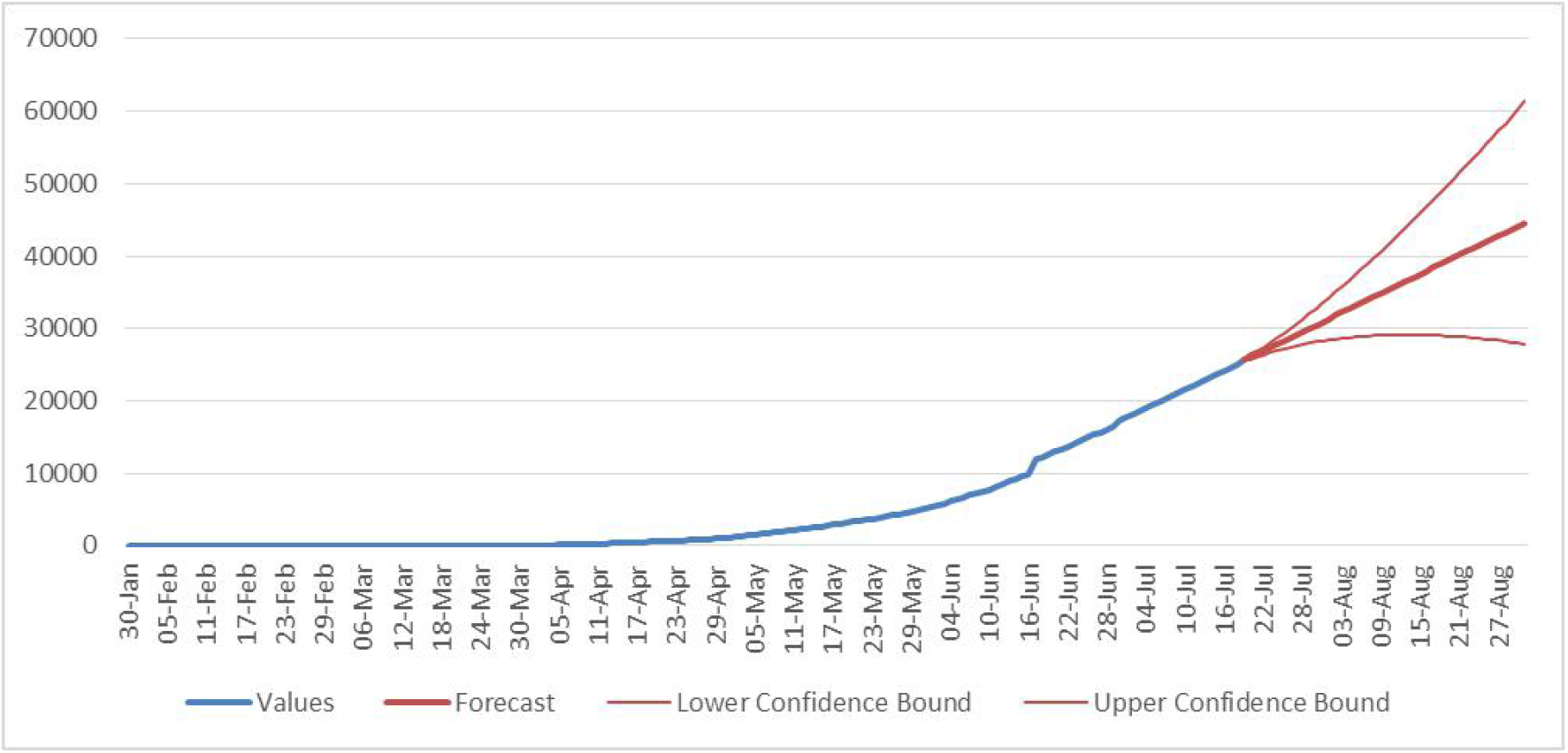
Forecasting for the death cases (ARIMA-311): India

### Holt-Winter method

The national statistics for COVID-19 spreading scenario for India up to 29^th^ June 2020 is shown in Figure 5. This figure shows the trend of the spread but, does not have seasonality. Hence, Holt-Winter model becomes Holt’s Method. To obtain the best time series forecasting model with COVID-19 dataset for India and its states, several attempts are made with different values of smoothing parameters α and β. The forecasting results using α=0.9, β=0.3 provides the best result for daily Confirmed cases for India and α=0.5, β=0.7 is best for predicting the daily death cases for India, with low AIC and RMSE. Further, forecasting results with a 95% confidence interval with upper bound and lower bound value estimation for confirmation and death cases are presented in Table 6. Finally, a comparison is made with actual COVID-19 spreading in India from 1^st^ July 2020 to 10^th^ July 2020, with the estimated values obtained from Holt-Winters model for the accuracy of the prediction, which is shown in Table 7.

**Table 6:**
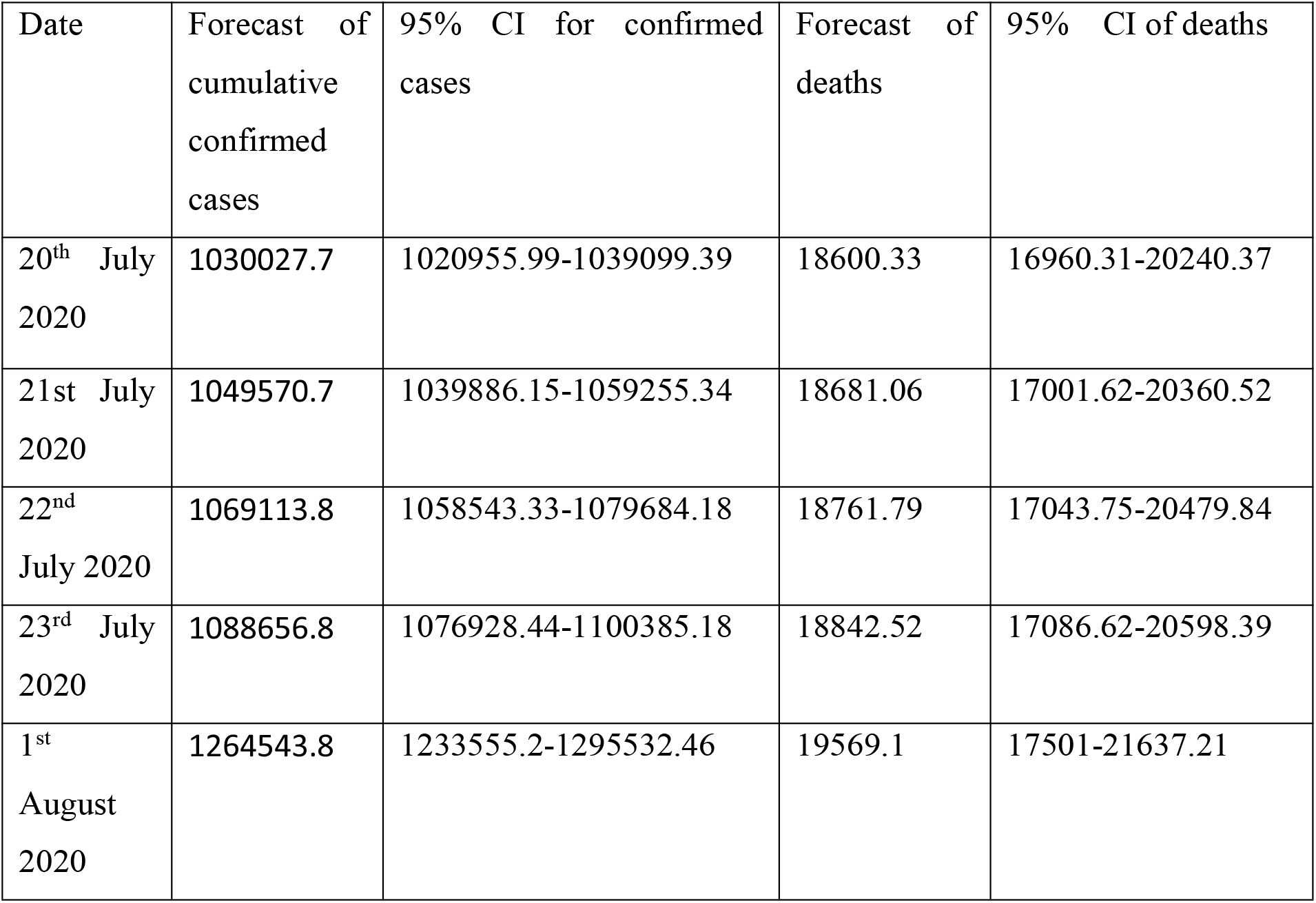
Results of 20-days ahead forecasting for confirmed and death cases using Holt-Winters model-India.

**Table 7:** comparison with official and estimated confirmed and death cases using Holt-Winters - India.

| date | official |  | Estimated |  | Accuracy % |  |
| --- | --- | --- | --- | --- | --- | --- |
|  | confirm | death | confirm | death | confirm | death |
| 1-7-2020 | 6,04,678 | 17840 | 677718.7 | 17066.461 | 87.92 | 95.66 |
| 2-7 | 6,25,020 | 17852 | 696062.8 | 17149.191 | 88.63 | 96.06 |
| 3-7 | 6,35,768 | 18219 | 714407 | 17227.922 | 87.63 | 94.56 |
| 4-7 | 670035 | 19259 | 732751.1 | 17308.652 | 90.63 | 89.87 |
| 5-7 | 696876 | 19693 | 751095.3 | 17389.382 | 92.21 | 88.30 |
| 6-7 | 716976 | 20151 | 769439.5 | 17470.113 | 92.68 | 86.69 |
| 7-7 | 739523 | 20623 | 787783.6 | 17550.843 | 93.47 | 85.10 |
| 8-7 | 764071 | 20642 | 806127.8 | 17631.574 | 94.49 | 85.41 |
| 9-7 | 794842 | 21623 | 824472 | 17712.304 | 96.27 | 81.91 |
| 10-7 | 819374 | 22107 | 842816.1 | 17793.034 | 97.13 | 80.48 |

**Fig 5:**
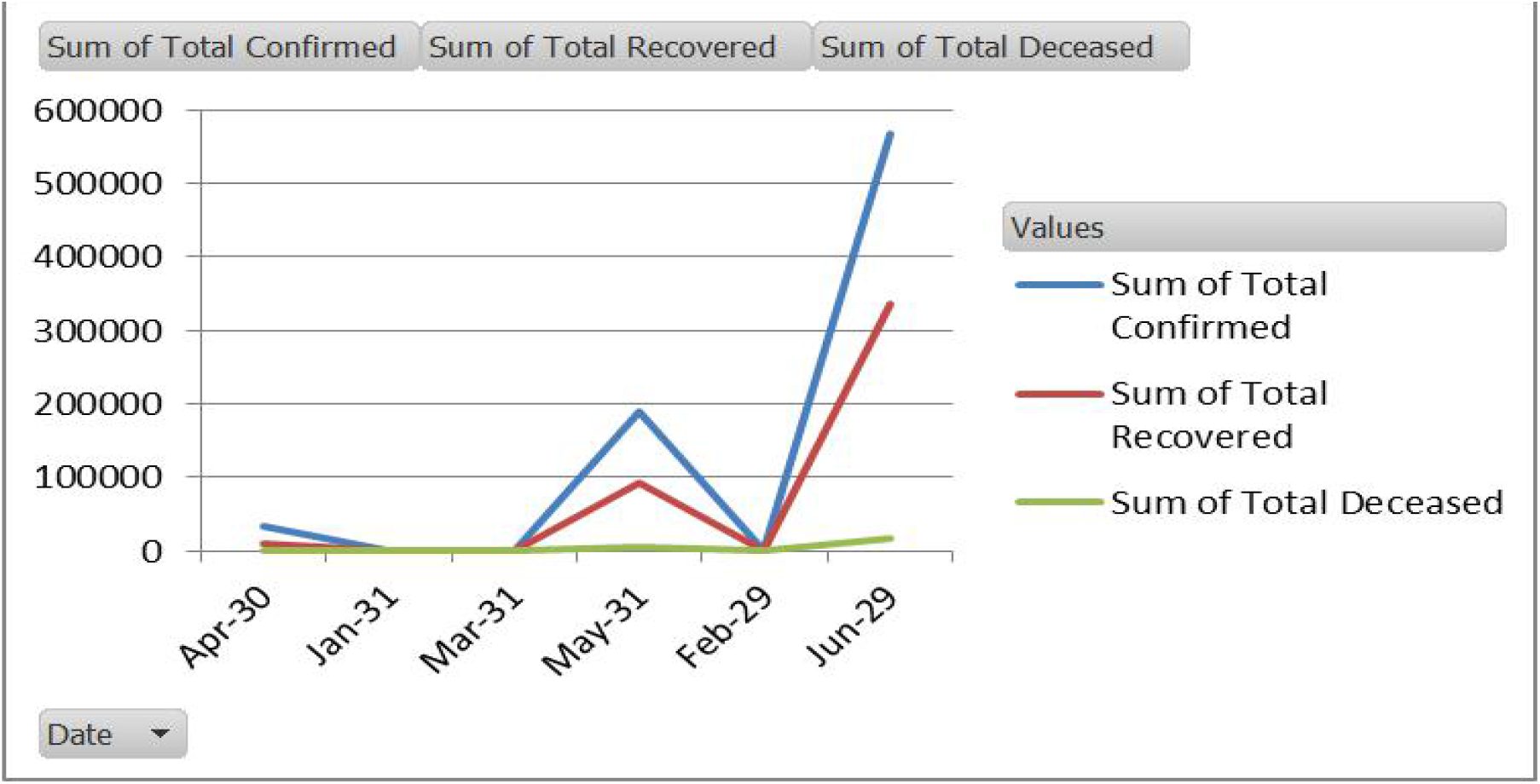
India statistics for COVID-19 spreading.

The proposed forecasting using Holt-winters model for confirm and death cases as India perspective are shown in Figure 6 and Figure 7 respectively.

**Figure 6:**
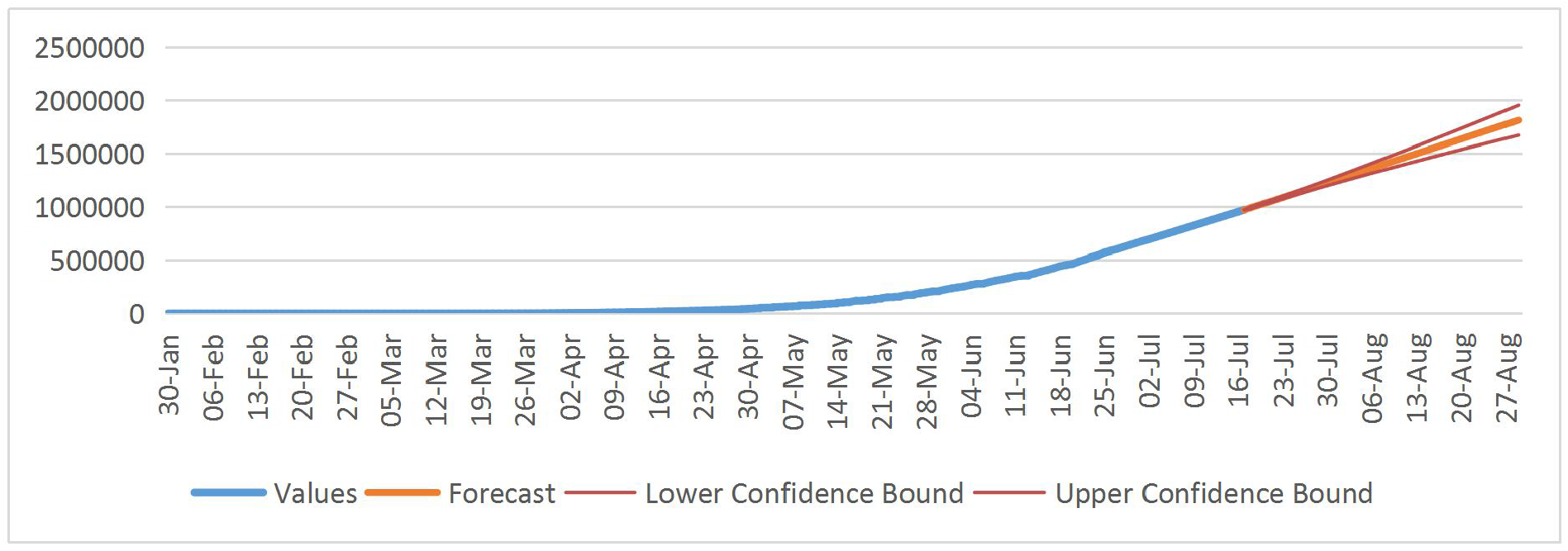
Holt-Winters forecasting for cumulative confirm cases in India

**Figure 7:**
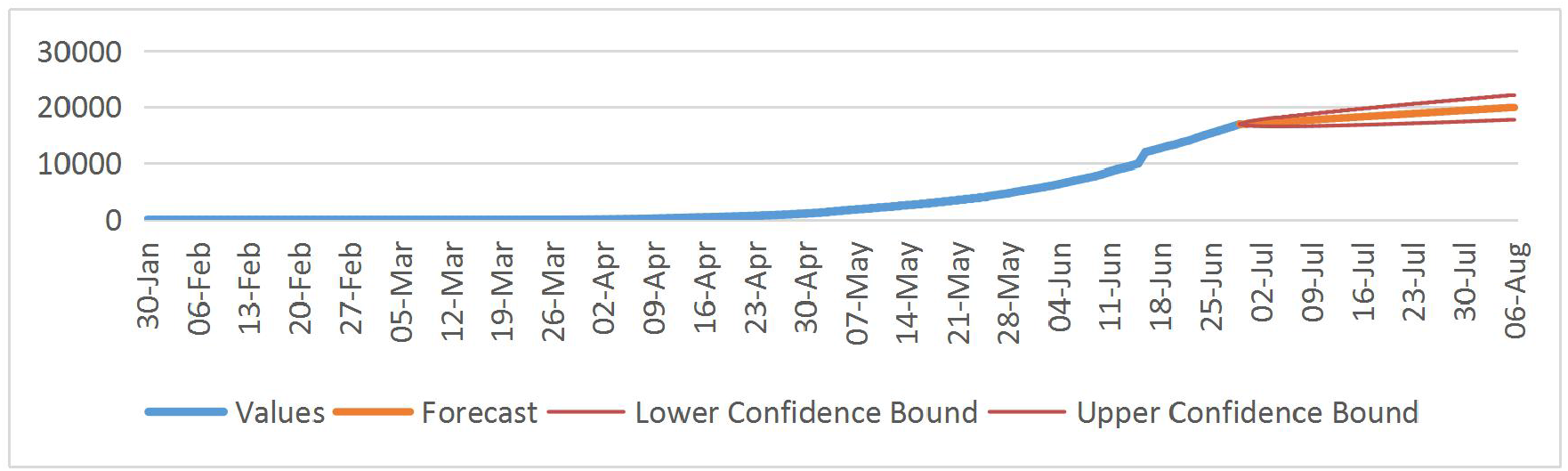
Holt-Winters forecasting for cumulative death cases in India

### Forecasting for Indian states

Looking into the fast trend of COVID-19 spread in India, investigating state-wise spreading of this contagious disease has become inevitable to contain the disease by taking some effective measures such as: making containment zones, stopping of the interstate bus and train services, effective lockdown and shutdown measures, building health care facilities etc. to name a few. Five Indian states: Maharashtra, Delhi, Andhra Pradesh, West Bengal and Odisha are selected for regional level short term forecasting using ARIMA and Holt-Winters model.

Figure 8 to Figure 12 shows the ARIMA forecasting with 411 and 311 for confirm and death cases, as they present the lowest AIC values and have p-value within the significance level.

**Figure 8:**
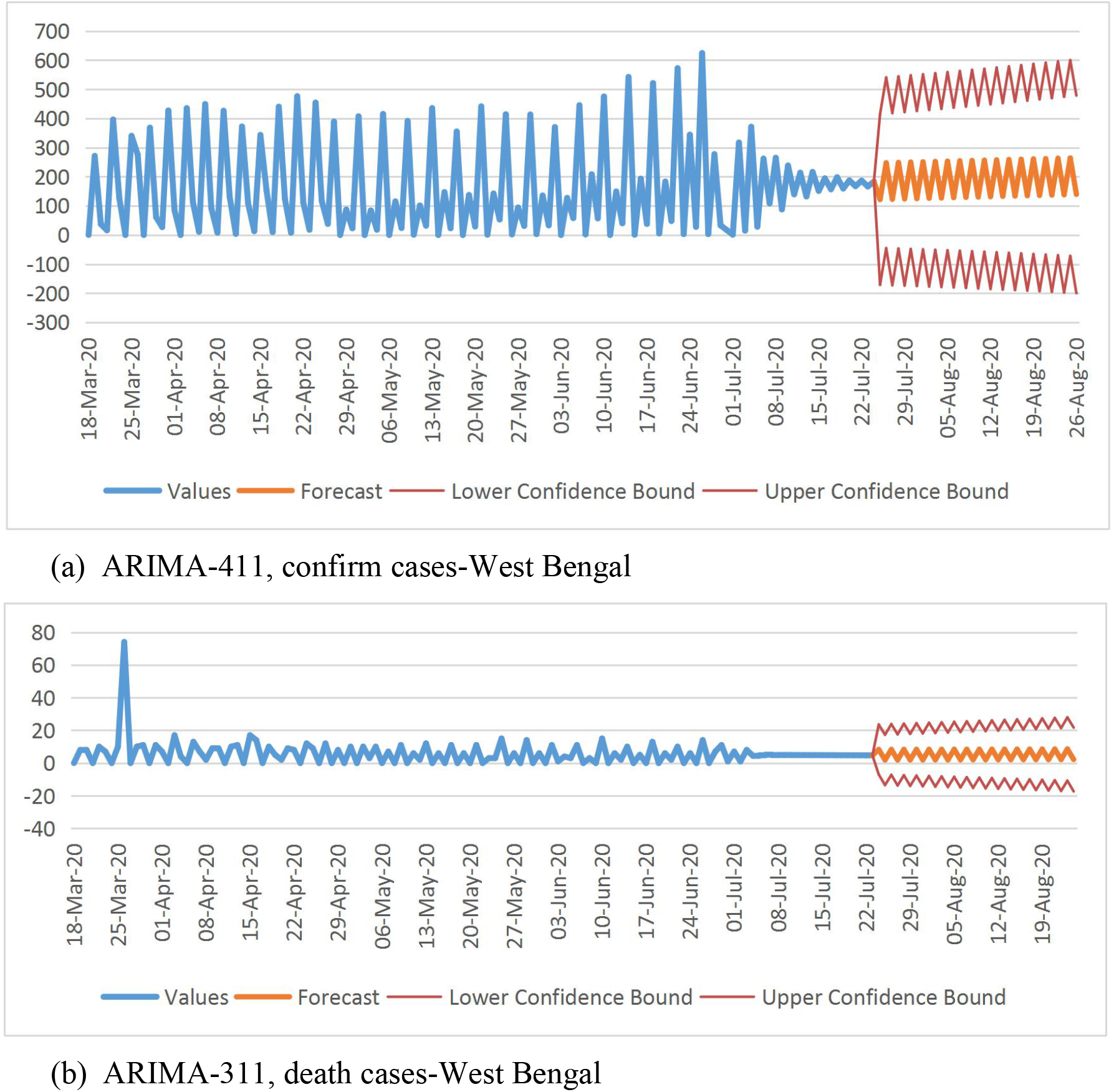
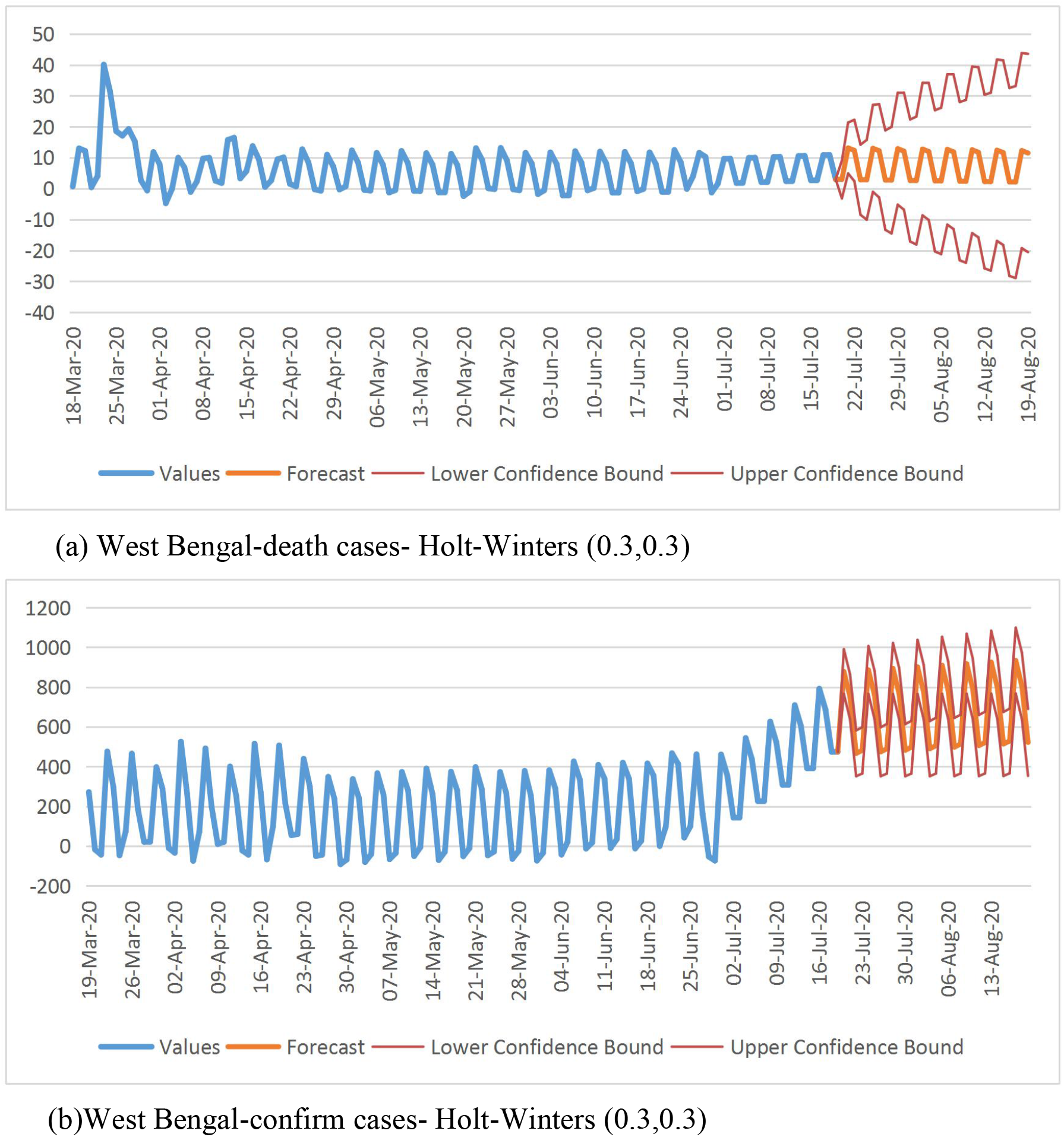
ARIMA and Holt-Winters forecasting for the confirmed cases and death cases for West Bengal

**Fig 9:**
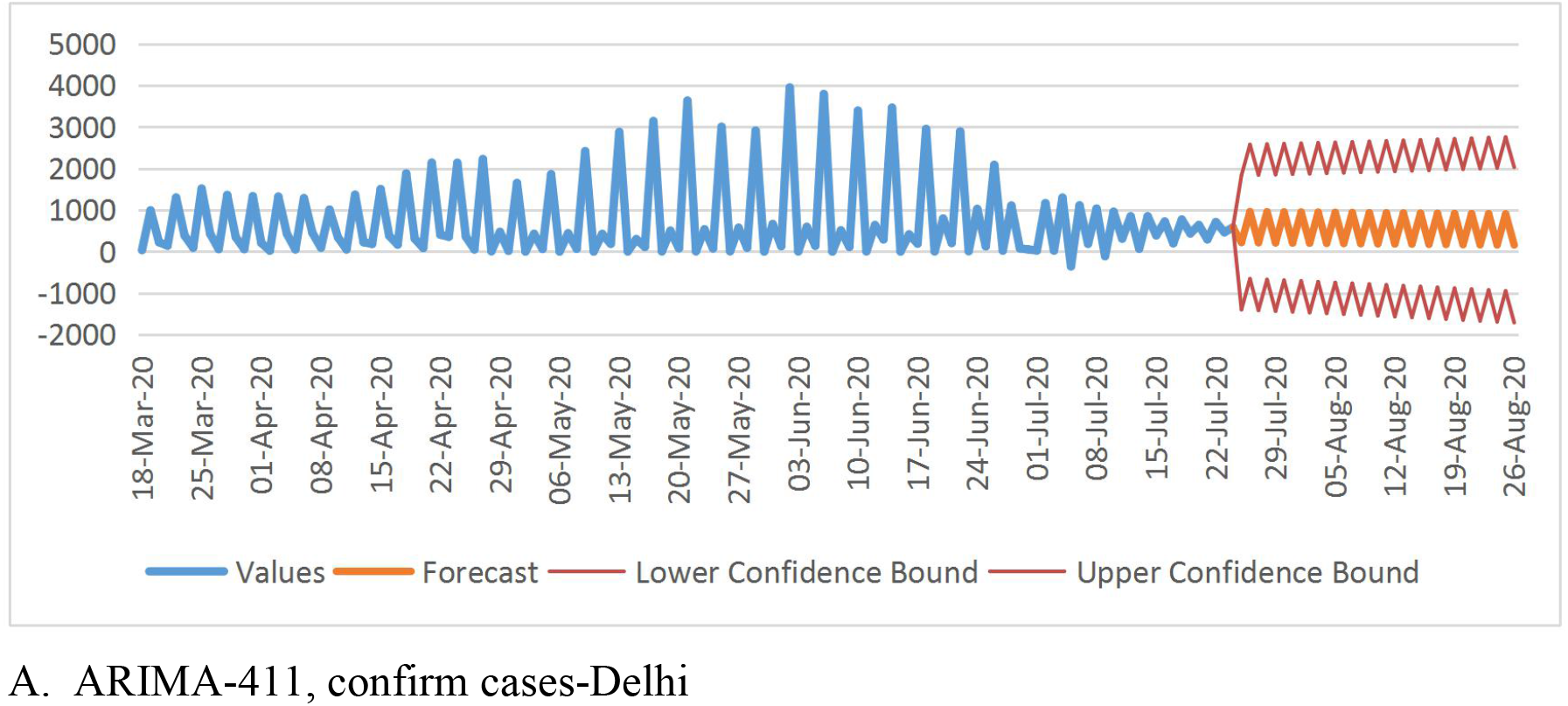
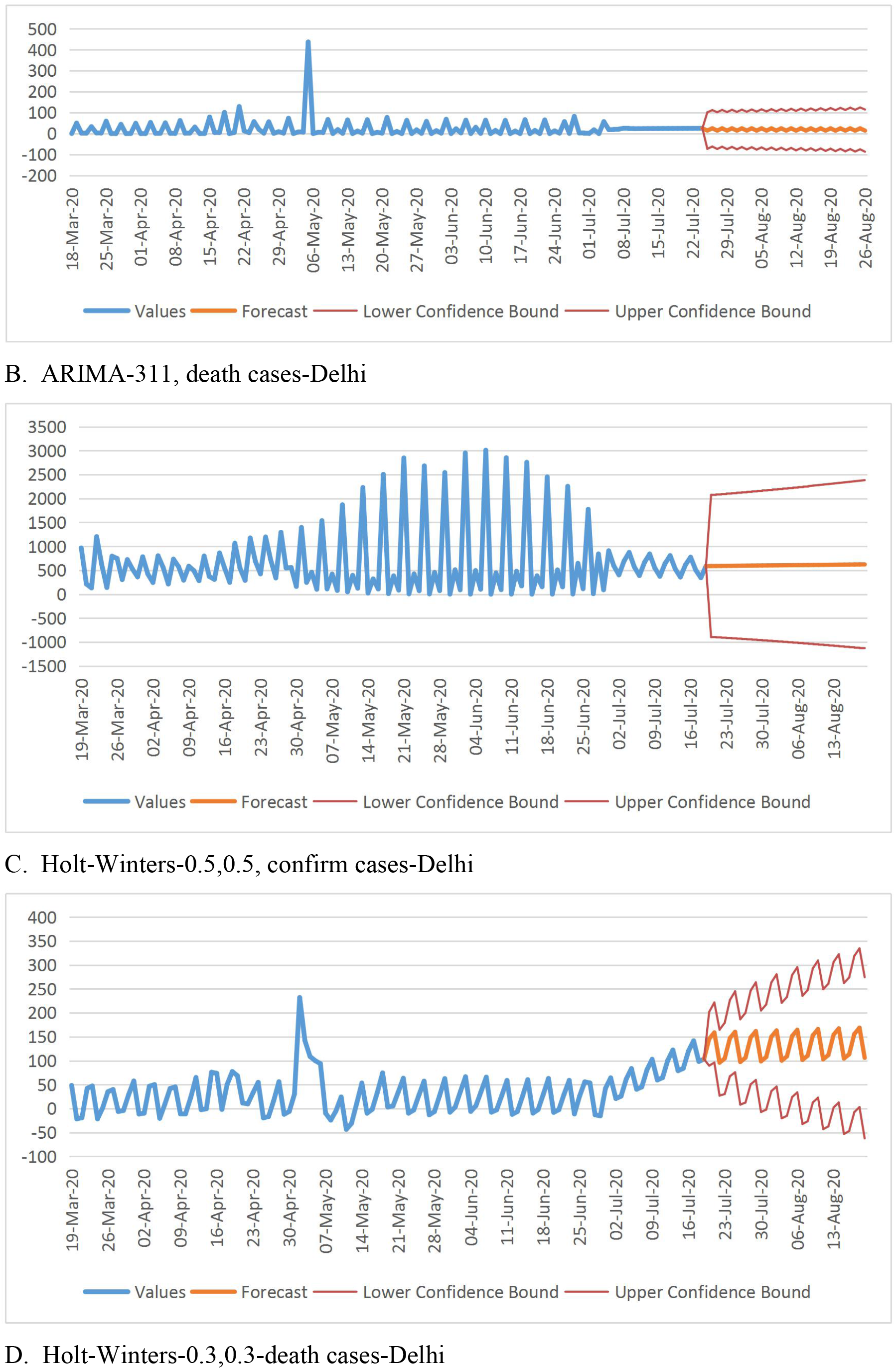
ARIMA and Holt-Winters forecasting for the confirmed cases and death cases For Delhi

**Fig. 10:**
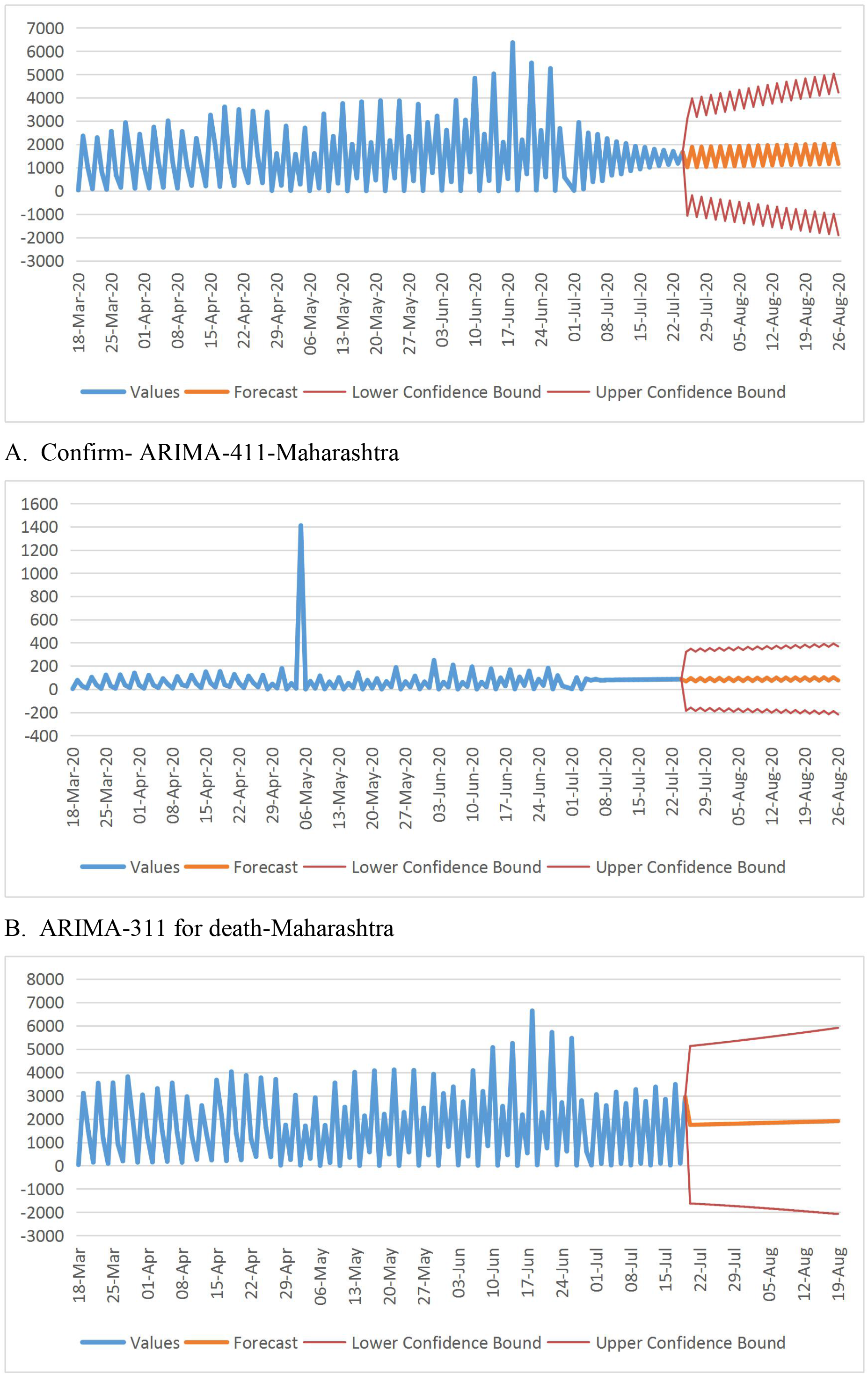
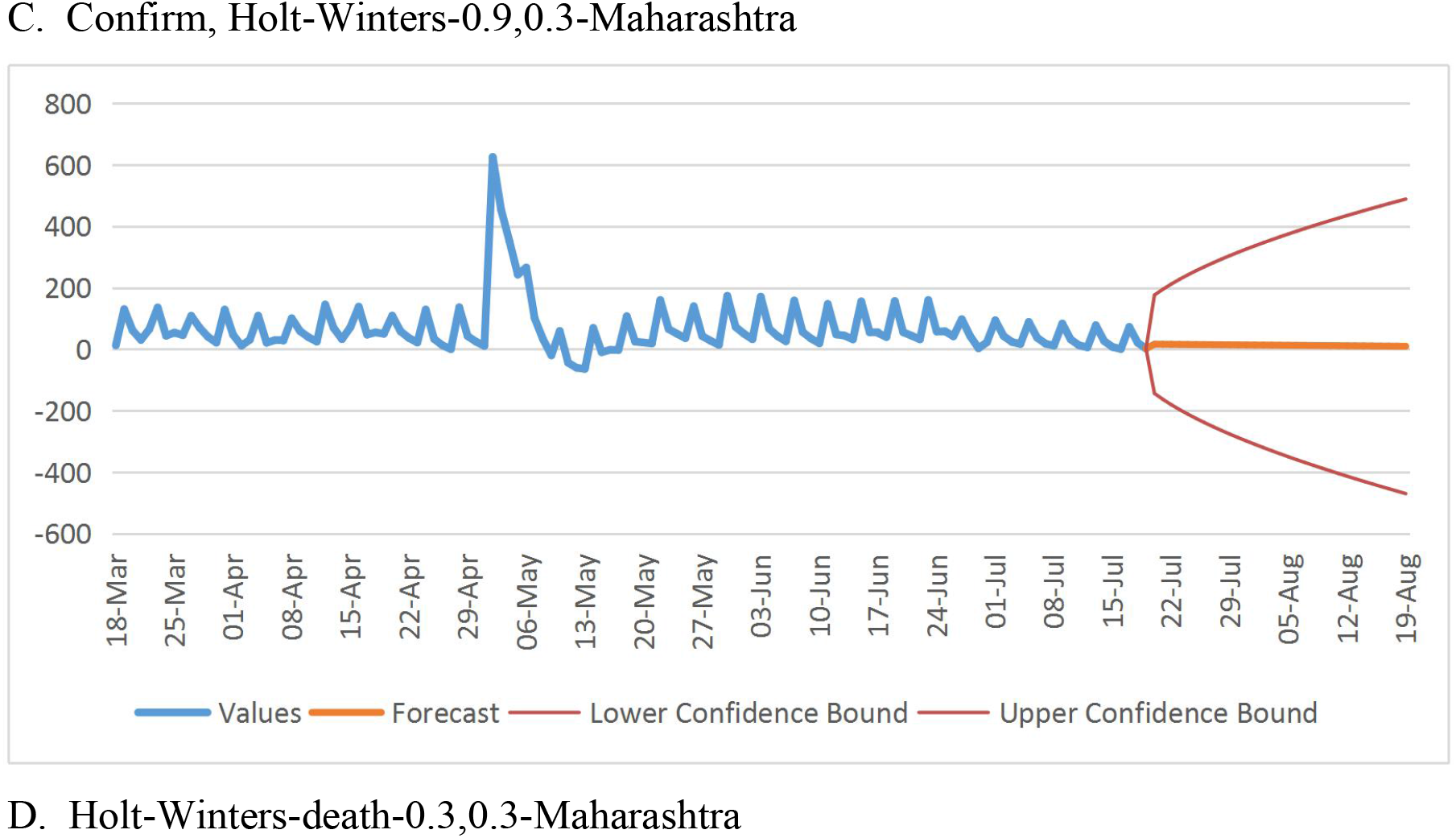
ARIMA and Holt-Winters forecasting for the confirmed cases and death cases For Maharashtra

**Fig 11:**
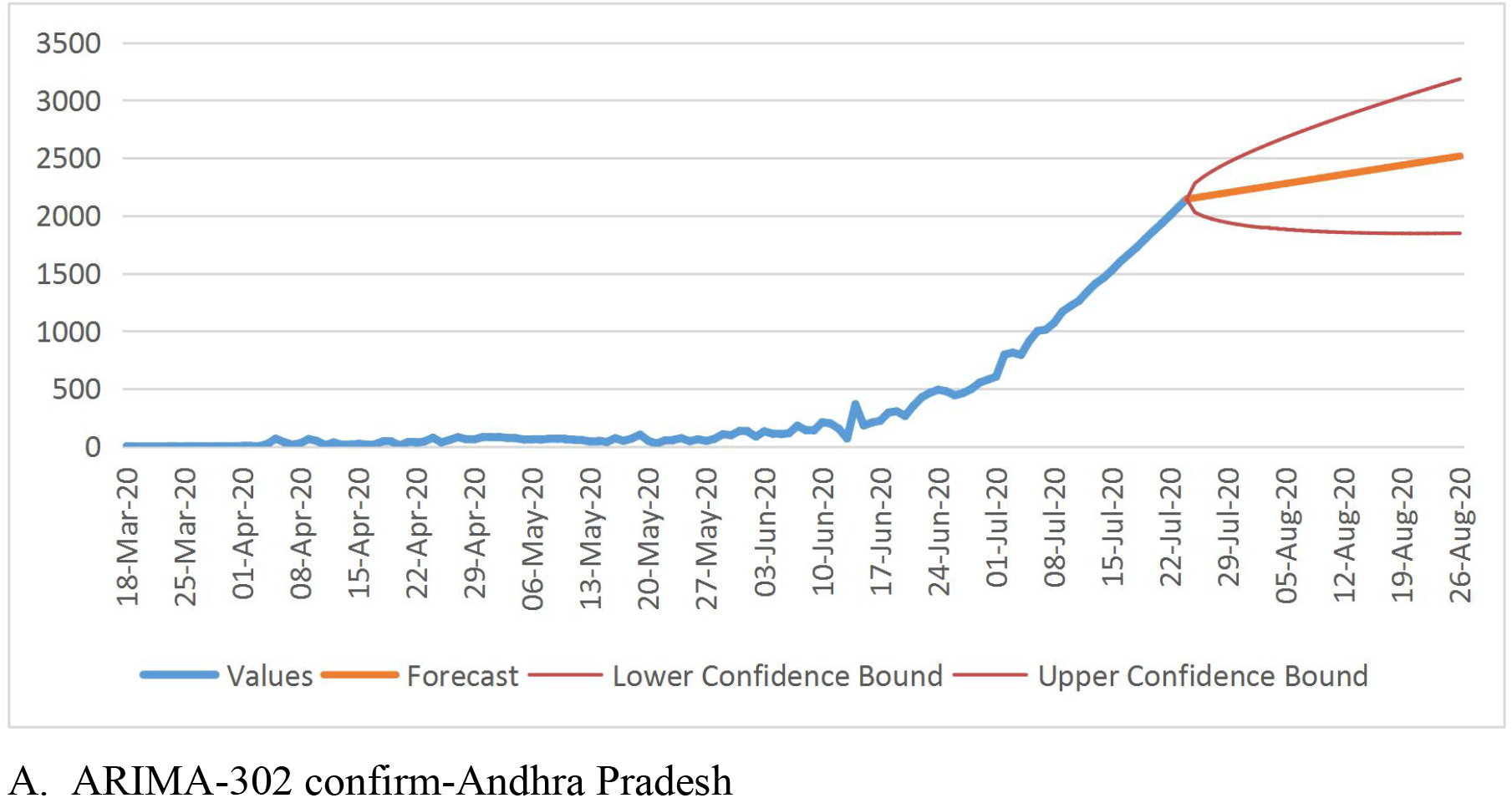
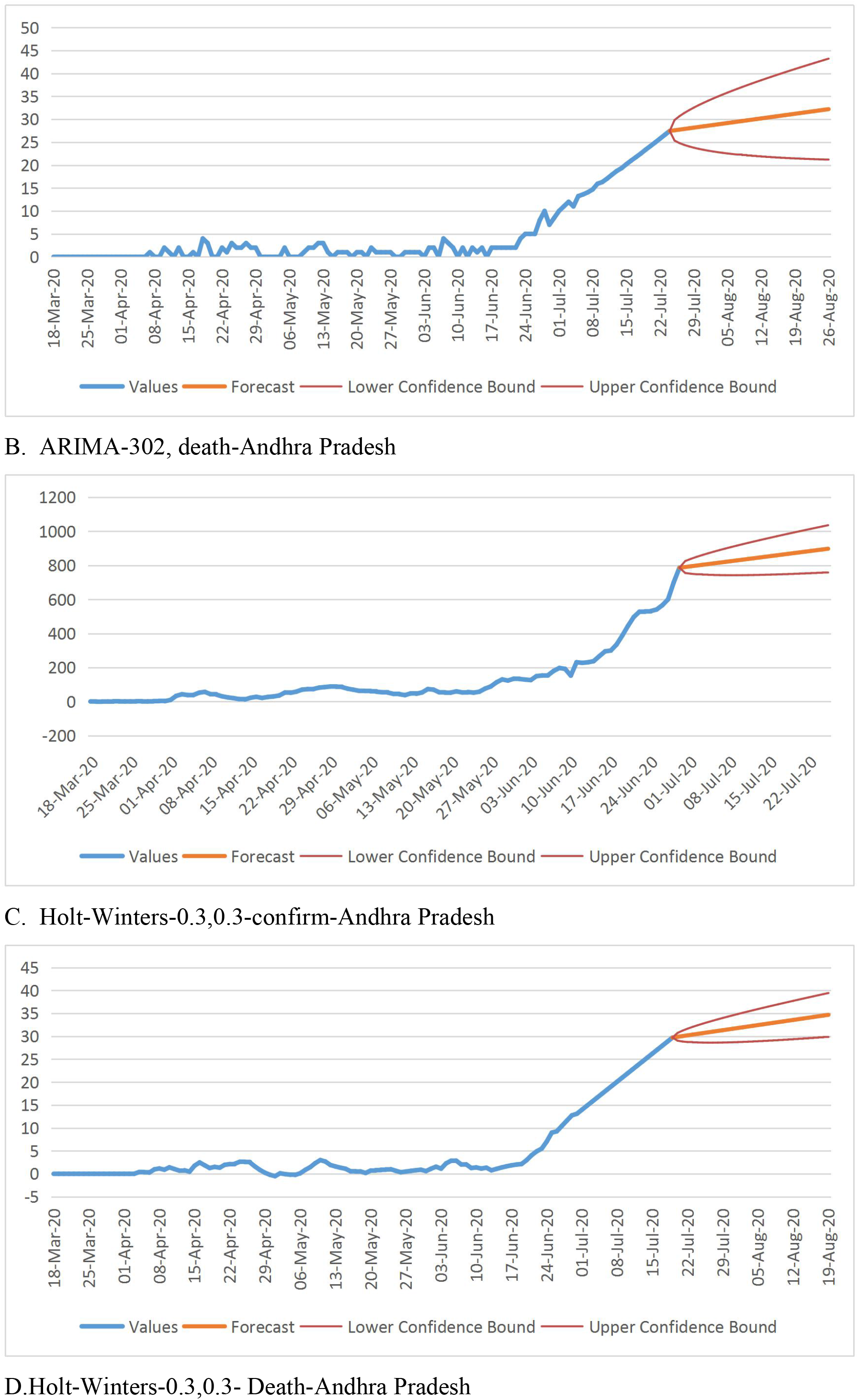
ARIMA and Holt-Winters forecasting for the confirmed cases and death cases For Andhra Pradesh

**Fig 12:**
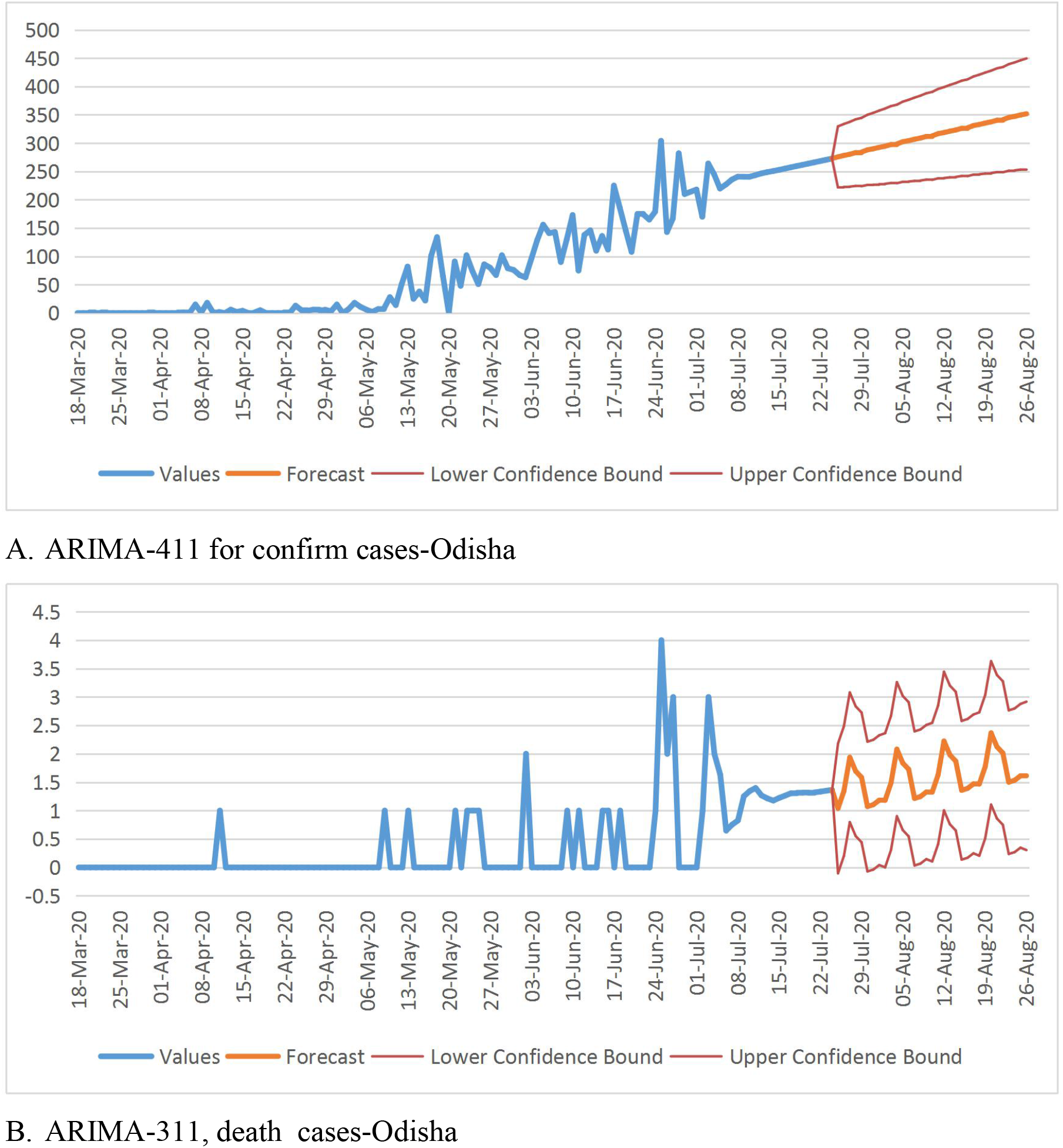
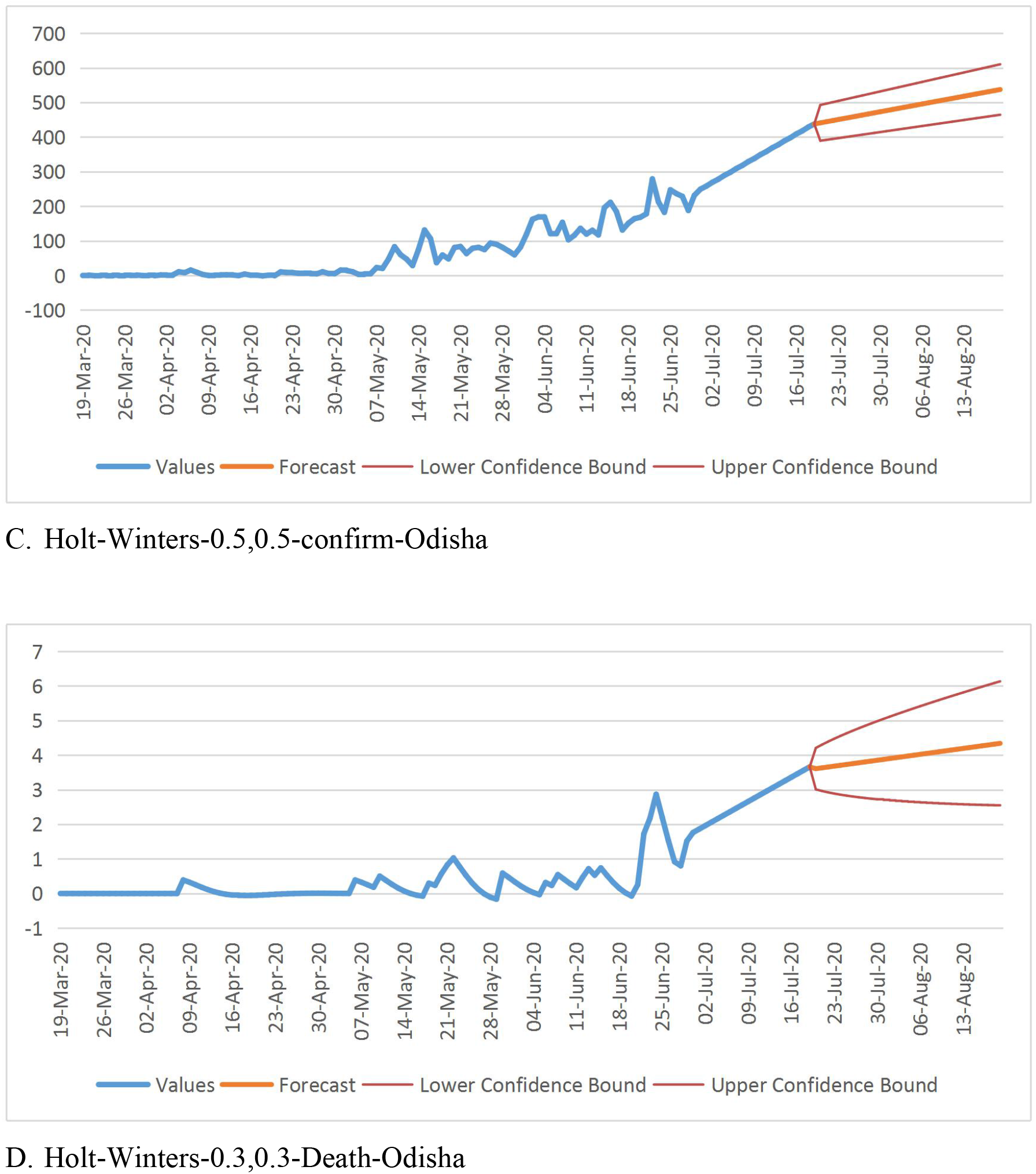
ARIMA and Holt-Winters forecasting for the confirmed cases and death cases for Odisha

The similar procedure adopted as done in case of India as a whole for forecasting, for the Indian states also. The AIC and RMSE value for the best chosen ARIMA model with point forecast and 95% confidence interval forecast with upper bound and lower bound values for both confirm and death cases are presented in Table 8.

**Table 8:**
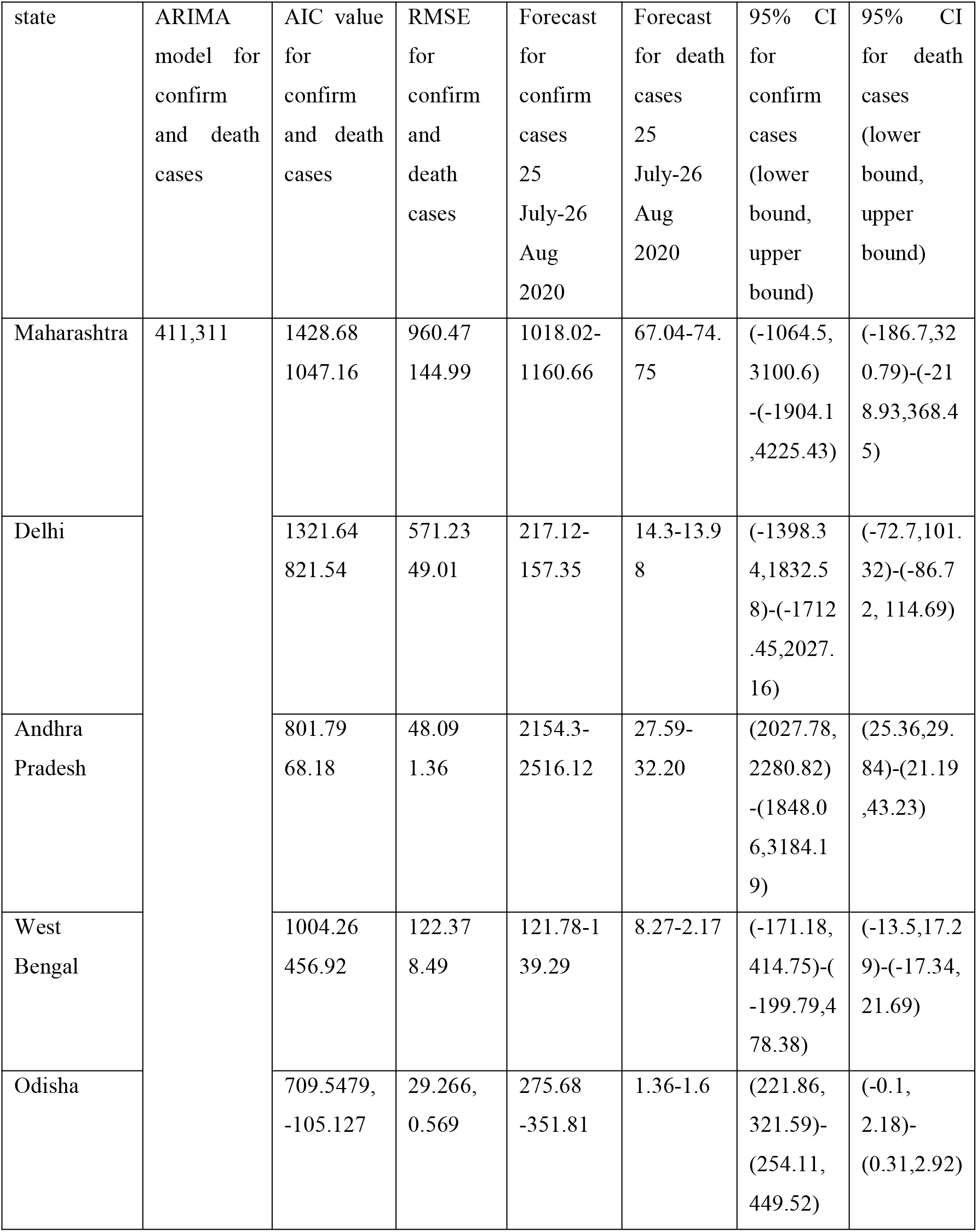
forecasting Indian states from 25^th^ July to 26^th^ August daily.

From Figure 8, it is understood that Maharashtra may have 3100 to 4225 daily confirmed cases with 1832 to 2027 for Delhi, 2280-3184 for Andhra Pradesh, 44-478 for West Bengal and 321-449 for Odisha. At the same time, the death cases for each state are 320-368 for Maharashtra, 101-114 for Delhi, 29-43 for Andhra Pradesh, 17-21 for West Bengal and 2-3 for Odisha. These forecast results show that Maharashtra is the hot spot for India with more number of confirmed and death cases till 26^th^ August 2020 and may continue further, unless some vaccines are discovered. Even though, Andhra Pradesh surpassed Delhi in daily confirmed cases, a big relief for the state is in having lower death in comparison to Delhi. For West Bengal and Odisha, even though confirmed cases have increased over the period but still it is under control in comparison to other Indian states. Odisha wins the race by having a very low daily death number limited to a maximum of 3.

All these results indicate that the COVID-19 pandemic to stay for a longer period and hence appropriate measures are sought to fight it successfully till some remedial measures are found to counter it.

## 5. Conclusions

ARIMA and Holt-Winters exponential smoothing methods are employed in this paper, to forecast and predict the model for 20-days ahead forecasting the COVID-19 cumulative confirmed, recovered and death cases as an Indian perspective from 30^th^ June to 19^th^ July 2020 and then, further extended up to 31^st^ August 2020. It is observed from the experiments that our proposed ARIMA model (411 for confirmed cases and 311 for death cases) is most accurate in forecasting the future cases with 99.8% and 99.3% respectively, in comparison to Holt-Winters method with 87.9% and 95.6%. The India forecasting reveals that the COVID-19 spreading will grow further in the long run and needs special attention by the people and the government to take precautionary measures to counter the disease effectively. As can be seen from the Indian state’s forecasting, there is a growing concern as the trend is increasing which was initially less in both confirmed and death cases for the states like Odisha. The forecast for India shows that there will be total 1886403.8 number of confirmed cases with lower and upper bound of 1728411.67-2044395.94 and 44603.10 with lower and upper bound of 27866.67-61339.84 death cases by 31^st^ August 2020. The forecasting results show that Maharashtra continues to be the hot spot for India with more number of deaths followed by Delhi. Odisha managed well by restricting both the total number of daily confirmed and death cases.

## Data Availability

MoHaFW daily data on COVID-19 
Full COVID-19 data for India obtained from kaggle

https://www.kaggle.com/imdevskp/covid-19-in-india

